# An Opportunity to Standardize and Enhance Intelligent Virtual Assistant-Delivered Layperson Cardiopulmonary Resuscitation Instructions

**DOI:** 10.1101/2023.03.09.23287050

**Authors:** William Murk, Eric Goralnick, John S. Brownstein, Adam B. Landman

## Abstract

**Importance:** Intelligent virtual assistants (IVAs) are ubiquitous and hold the potential to provide bystander cardiopulmonary resuscitation (CPR) instructions during an emergency.

**Objective:** To evaluate the quality of CPR instructions provided by IVAs.

**Design:** We evaluated the appropriateness of responses of four IVAs (Amazon Alexa, Apple Siri, Google Assistant, and Microsoft Cortana) to eight CPR-related questions. We also evaluated text-based responses provided by OpenAI ChatGPT, a recently developed artificial intelligence large language model.

**Results:** Out of 32 responses provided by IVAs, only 19 (59%) were related to CPR, 9 (28%) suggested calling emergency services, and 4 (12%) provided verbal CPR instructions. All responses provided by ChatGPT were related to CPR and suggested calling emergency services. Among responses related to CPR, the answers provided varied significantly in the utility of information provided.

**Conclusions and Relevance:** These results highlight the need for the technology industry to partner with the medical community to improve and standardize bystander CPR instruction provided by IVAs.

**Key points:** *Question:* What is the quality of cardiopulmonary resuscitation (CPR) instruction provided by intelligent virtual assistants (IVAs) including Amazon Alexa, Apple Siri, Google Assistant, and Microsoft Cortana?

*Findings:* When asked questions related to CPR, IVAs frequently did not provide relevant information. Moreover, even when the information provided was relevant, actual CPR instruction was not given in most cases.

*Meaning:* These results highlight the need to standardize bystander CPR instruction provided by IVAs.

## Introduction

More than 23.8 million viewers watched Damar Hamlin’s cardiac arrest and resuscitation live on “Monday Night Football”, providing an unprecedented opportunity for engaging the public on cardiopulmonary resuscitation (CPR) and automated external defibrillator (AED) use.^1^ Despite widespread CPR courses and public campaigns, only 46% of out of hospital cardiac arrest (OHCA) cases have layperson CPR performed.^2^ Each minute a patient is in OHCA without cardiopulmonary resuscitation (CPR) or defibrillation decreases survival by 7-10%.^3^ Layperson CPR can increase survival by 2-4 fold.^3^

Intelligent virtual assistants (IVAs) are becoming ubiquitous and may be a novel method to provide CPR instructions to bystanders during an emergency. IVAs respond to verbal queries using speech recognition and other forms of artificial intelligence. These technologies are used by nearly half of US adults^4^ and are available on standalone smart speakers, like Amazon Echo or Google Nest, or integrated into smartphones and computers, such as Apple Siri on iOS/Mac devices. IVAs are increasingly being used for healthcare needs, with 8% and 21% of US adults reporting such use in 2019 and 2021, respectively.^5^

Although bystanders may obtain CPR instructions from emergency dispatchers, such services are not universally available, and their use may be limited or delayed by language barriers, poor audio quality, call disconnection, fear of law enforcement, and perceived costs of using emergency services.^6^ IVAs may therefore serve as the primary source of readily accessible hands free CPR instruction in the bystander’s preferred language when it otherwise may not be promptly accessed.

Given their potential to improve public health, we tested common IVAs for their out of the box ability to provide appropriate CPR instruction. Four IVAs were tested, including Amazon Alexa, Apple Siri, Google Assistant, and Microsoft Cortana, and their responses to eight CPR-related questions were evaluated. To assess whether responses improved over time, we compared the quality of responses between 2022 and 2023. In addition, as a secondary objective, we tested the ability of ChatGPT, a recently developed artificial intelligence large language model, for its ability to provide CPR instruction. Although ChatGPT, in its native format, only responds to text-based queries and provides text-based answers and is therefore not an IVA, we sought to evaluate its performance as it may be indicative of future IVA capabilities.

## Methods

We tested several ways people might ask about CPR on four popular IVAs (Amazon Alexa on Echo Show 5, Apple Siri on iPhone X [iOS 15.3.1] in Feb. 2022 and iPhone 14 Pro [iOS 16.2] in Mar. 2023, Google Assistant on Nest Mini, and Microsoft Cortana on a Windows 10 [version 21H2] laptop). Testing was conducted in March 2022 and February 2023, and the results were compared across time periods. Eight verbal queries that could indicate a need for CPR instruction were asked of each IVA. To ensure that responses were not dependent on accuracy of voice recognition, text-based transcriptions of interactions (as provided by all four IVAs) were reviewed to verify that verbal queries were accurately captured. Only responses immediately provided by the IVA (whether verbal or textual) were evaluated; if a response included a link to a website providing further information, that website was not included for evaluation. In addition, we also asked the eight queries to ChatGPT (OpenAI, Feb 13 Version). Since ChatGPT, in its native format, does not provide verbal interaction, these queries and responses were text-based. For ChatGPT, queries were assessed in February 2023, and a new chat was started for each query to prevent prior queries affecting the responses provided.

All queries were made while geolocated in the United States. Two board-certified emergency medicine physicians (EG and ABL) rated whether each response was related to CPR, whether emergency medical services (EMS) were recommended, whether any CPR instructions were provided (verbal or textual), and whether verbal CPR instructions were provided. There are no human subjects involved in this work and therefore we did not pursue Institutional Review Board review or informed consent.

## Results

**Figure 1** illustrates responses to CPR queries by IVA in 2023. Out of 32 responses, 19 (59%) were related to CPR, 7 (22%) were unrelated, and 6 (19%) acknowledged that the answer was not known. Only 9 (28%) suggested calling emergency services, 11 (34%) provided any (verbal or textual) CPR instructions, and 4 (12%) provided verbal CPR instructions (**Figure 2**). These responses were only modestly improved compared to responses from the previous year (**Figure 2**). Differences in appropriateness of responses were noted across IVA products. For example, while Microsoft Cortana more frequently provided CPR instructions than other IVAs, these instructions were textual only, and it did not provide any verbal instructions (**Figure 3**). Full transcriptions of responses are provided in **eTable 1 - 4**.

**Figure 1.**
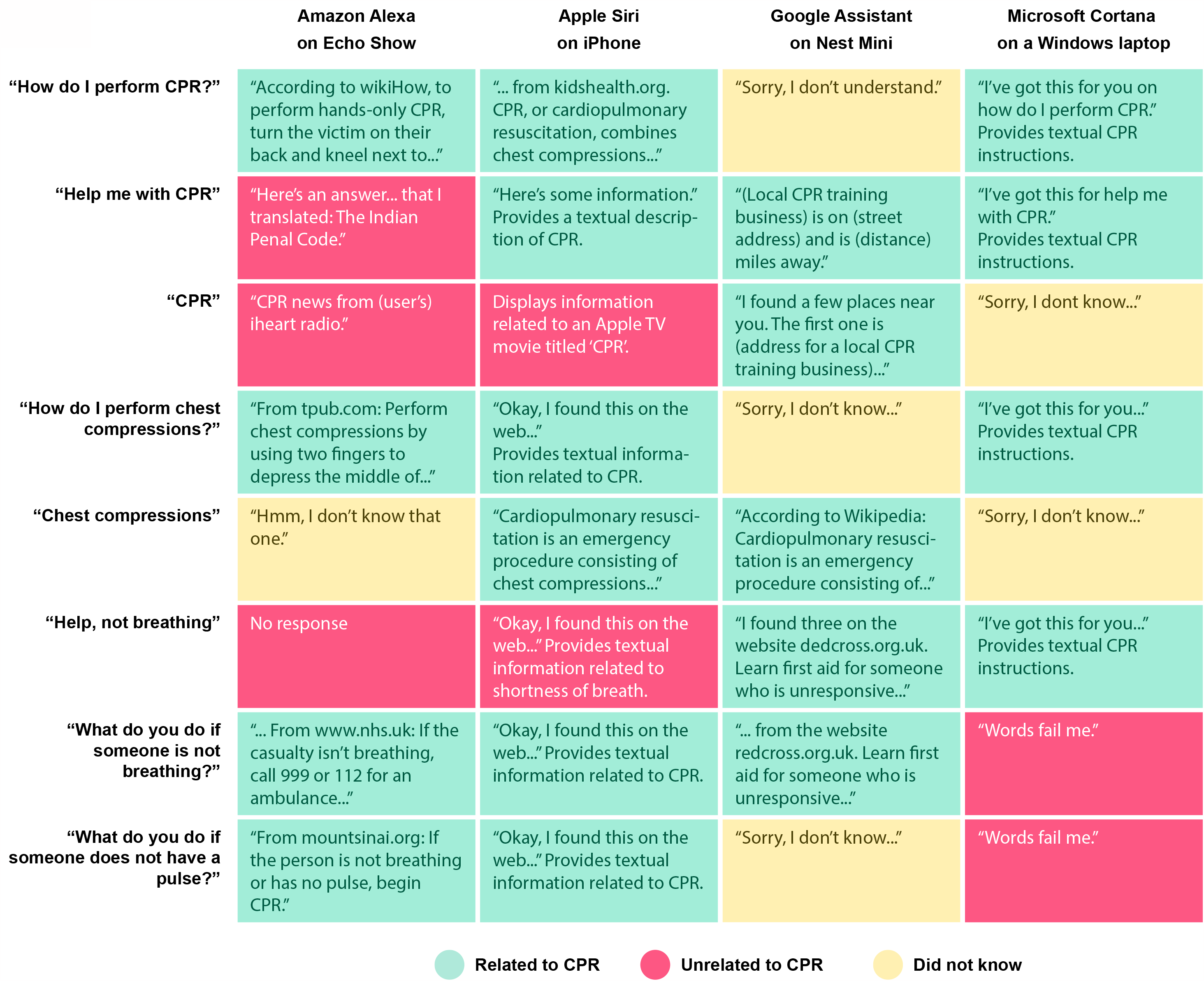
Responses to CPR questions, by intelligent virtual assistant (IVA) in February 2023. Responses are colored according to whether the response was determined to be related to CPR (providing information pertaining to CPR or recommending the use of emergency services), unrelated, or the IVA acknowledged that it did not know the answer. Responses shown here are abbreviated versions of the full response transcriptions provided in eTables 1-4.

**Figure 2.**
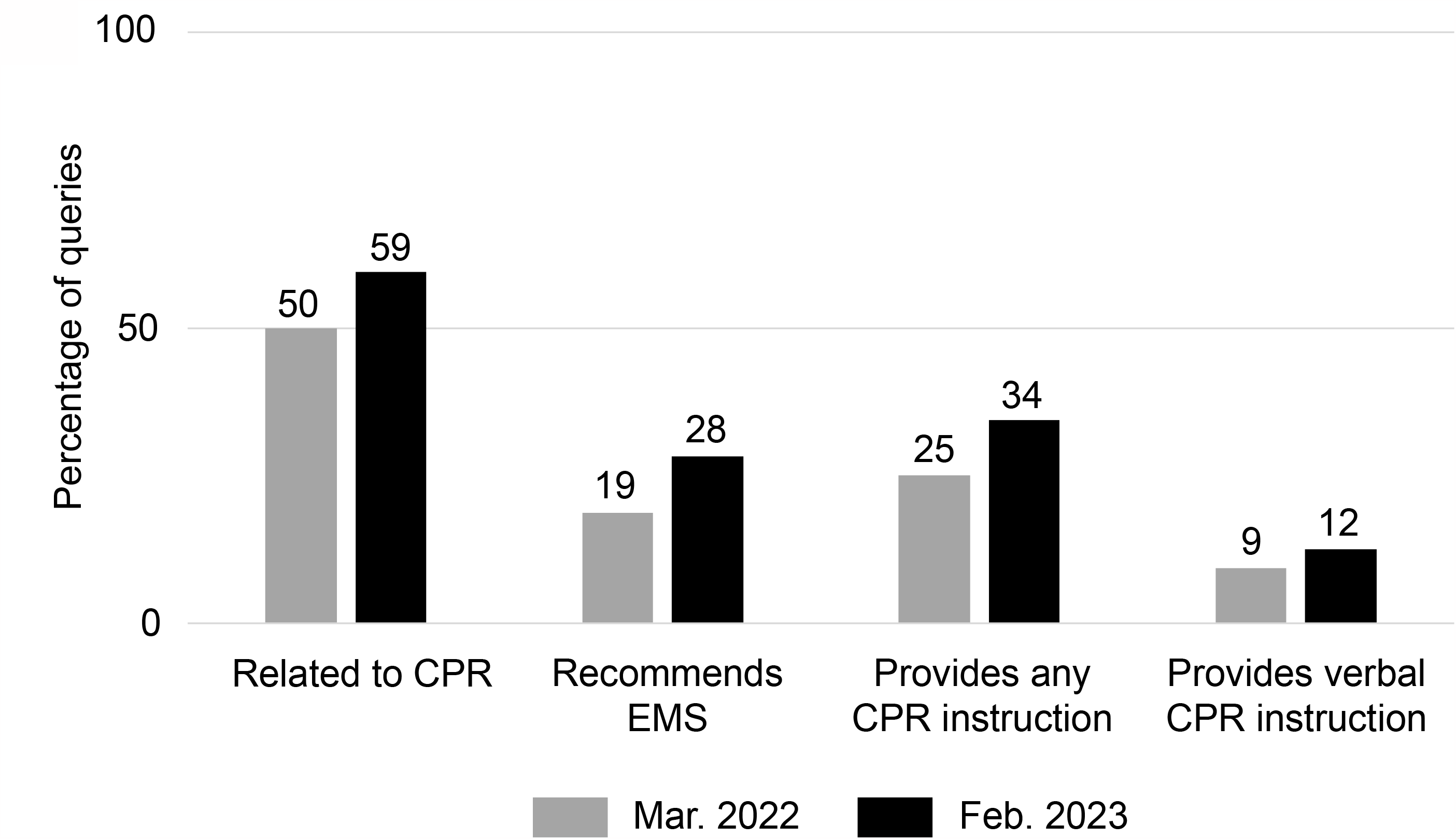
Summary of responses to CPR questions, across all four IVAs, in March 2022 and February 2023. N = 32 questions. ChatGPT results not included.

**Figure 3.**
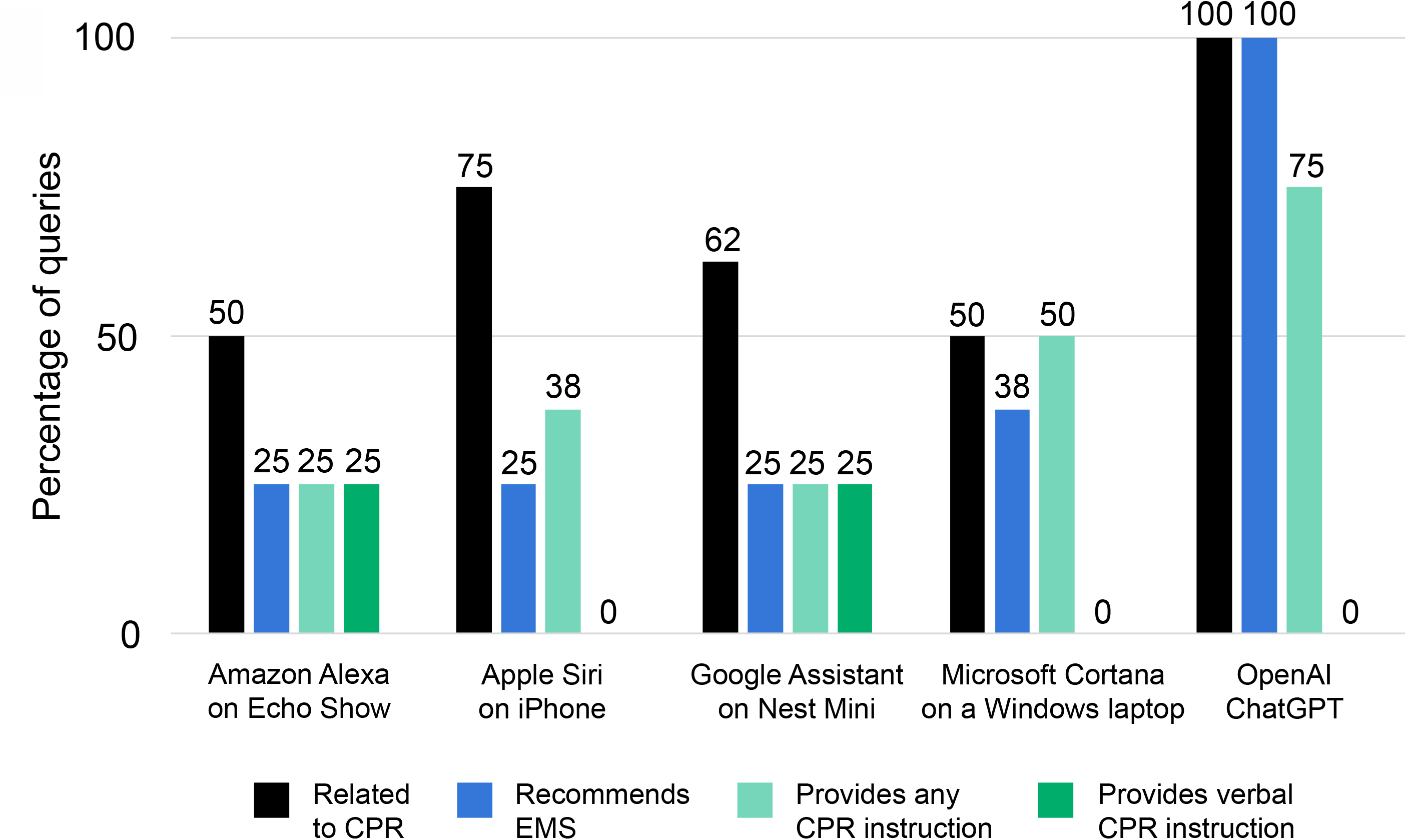
Summary of responses to CPR questions, by product, in February 2023. N = 8 questions.

ChatGPT provided relevant CPR information and recommended emergency services for 100% of queries asked of it, and provided textual CPR instructions for 75% of queries (**Figure 3** and **eTable 5**).

Among the 17 responses from the IVAs or ChatGPT that provided any CPR instruction, there was considerable variability in the content of instruction provided. For example, only 12 (71%) described hand positioning, 8 (47%) described compression depth, 6 (35%) described compression rate, and 2 (12%) mentioned use of an AED (**eTable 6**). While most descriptions of compression depth recommended a depth of 2 inches, one response recommended a depth of 0.5 to 1 inch, suggesting that this may have been intended for infant CPR (**eTable 1A**).

Among responses recommending calling emergency services, some suggested calling 911, while others suggested calling 999 or 112, indicating that these products do not necessarily take geographic context into consideration when providing responses.

## Discussion

Among IVAs, nearly half of queries were answered with information unrelated to CPR, often leading to grossly inappropriate responses, such as recommending a radio station. Among CPR-related responses, the content of answers varied considerably, and in many cases the user must take an extra step of visiting a website to obtain CPR instructions. A well-intentioned layperson seeking to use an IVA for real-time CPR guidance may experience delays or not find appropriate content, which could worsen the patient’s condition. IVA performance has not substantially improved over time. Although ChatGPT had significantly improved performance compared to the IVAs, its responses nonetheless were inconsistent.

IVAs need to better support CPR by: 1) Building CPR instructions into core functionality (without requiring supplemental downloads); 2) Designating common phrases to activate CPR instructions; and 3) Establishing a single set of evidence-based content across devices. Interactivity may be required to provide suitable CPR instruction, such as prompting the user for the age category of the patient.

A 2020 study similarly found that IVAs provided generally poor first aid advice, suggesting that there has been little progress to take advantage of these technologies for empowering bystanders to provide medical care.^6^ There is ample opportunity for the technology industry to partner with the medical community and professional societies to improve public health. Standardizing and improving IVA support for bystander CPR instruction has the potential to improve OHCA survival. Furthermore, IVAs may be able to support layperson empowerment to perform key skills in other time-sensitive medical conditions including, for example, bleeding control, opiate overdose reversal, or use of AEDs.

## Data Availability

All data produced in the present work are contained in the manuscript

## Acknowledgements

We thank the late Walter L. Rosenzweig for inspiring the idea that virtual assistants could deliver real-time CPR instruction to bystanders and help save lives. We also thank Melissa Landman for her thoughtful review of the manuscript draft.

