## Supplement for "An Opportunity to Standardize and Enhance Intelligent Virtual Assistant-Delivered Layperson Cardiopulmonary Resuscitation Instructions"

**Table of contents**

**eTable 1A. Amazon Alexa on Echo Show: Responses to CPR questions. 2**
**eTable 1B. Amazon Alexa on Echo Show: Categorization of Responses. 3**
**eTable 3A. Google Assistant on Nest Mini: Responses to CPR questions. 6**
**eTable 3B. Google Assistant on Nest Mini:: Categorization of Responses. 7**
**eTable 4A. Microsoft Cortana on a Windows laptop: Responses to CPR questions. 8**
**eTable 4B. Microsoft Cortana on a Windows laptop: Categorization of Responses. 8**
**eTable 5A. OpenAI ChatGPT: Responses to CPR questions. 10**
**eTable 5B. OpenAI ChatGPT: Categorization of Responses. 12**
**eTable 6. Content of responses providing CPR instructions (Feb. 2023). 13**

### eTable 1A. Amazon Alexa on Echo Show: Responses to CPR questions.

Quotations indicate verbal responses. Gray text describes non-verbal responses. *Italics* represent contextual/explanatory information.

| Question | Response from March 2022 | Response from February 2023 |
|---|---|---|
| How do I perform CPR? | "According to wikiHow, to perform hands-only CPR, turn the victim on their back and kneel next to them. Place your hands on their chest, one on top of the other, and perform chest compressions by pressing down firmly and quickly on their chest, aiming to push down 2 inches each time. Call the emergency services and aim to do 100 compressions per minute until they arrive. To learn conventional CPR, including opening the victim's airway and performing rescue breathing, sign up for a class at an emergency training school in your area." | "According to wikiHow, to perform hands-only CPR, turn the victim on their back and kneel next to them. Place your hands on their chest, one on top of the other, and perform chest compressions by pressing down firmly and quickly on their chest, aiming to push down 2 inches each time. Call the emergency services and aim to do 100 compressions per minute until they arrive. To learn conventional CPR, including opening the victim's airways and performing rescue breaths, sign up for a class at an emergency training school in your area." |
| Help me with CPR | *Plays an audio tone only; no information displayed* | "Here is an answer from an Alexa Answers contributor that I translated: The Indian Penal Code." |
| CPR | "Should I play the station CPR?" | "CPR news from (User's) iheart radio." |
| How do I perform chest compressions? | "Here's something I found on the web. According to riversideonline.com: To perform chest compressions, kneel next to the person's neck and shoulders." | "From tpub.com: Perform chest compressions by using two fingers to depress the middle of the sternum approximately 1/2 to 1 inch." |
| Chest compressions | "Sorry, I don't know that one." | "Hmm, I don't know that one." |
| Help, not breathing | "If this is a life-threatening situation, please dial emergency services directly from your phone." | *No response* |
| What do you do if someone is not breathing? | "Here's something I found on the web. According to freepatentsonline.com: If the patient is not breathing, controlled ventilation should be provided." | "Here is what I found: From www.nhs.uk: If the casualty isn't breathing, call 999 or 112 for an ambulance and then begin CPR." |
| What do you do if someone does not have a pulse? | "Here's something I found on the web. According to patsnap.com: If the pulse is found to be not valid, processing continues in step 302 where the next pulse is measured." | "From mountsinai.org: If the person is not breathing or has no pulse, begin CPR." |

**eTable 1B. Amazon Alexa on Echo Show: Categorization of Responses.**

Green: relevant or yes; red: not relevant or no; yellow: voice assistant did not know.

| | Response from March 2022 | | | | Response from February 2023 | | | |
|---|---|---|---|---|---|---|---|---|
| | Relatedness to CPR | Recommends emergency services? | Any CPR instructions (text or verbal) | Verbal CPR instructions? | Relatedness to CPR | Recommends emergency services? | Any CPR instructions (text or verbal) | Verbal CPR instructions? |
| How do I perform CPR? | Related | Yes | Yes | Yes | Related | Yes | Yes | Yes |
| Help me with CPR | Unrelated | No | No | No | Unrelated | No | No | No |
| CPR | Unrelated | No | No | No | Unrelated | No | No | No |
| How do I perform chest compressions? | Related | No | Yes | Yes | Related | No | Yes | Yes |
| Chest compressions | Don’t know | No | No | No | Don’t know | No | No | No |
| Help, not breathing | Related | Yes | No | No | Unrelated | No | No | No |
| What do you do if someone is not breathing? | Unrelated | No | No | No | Related | Yes | No | No |
| What do you do if someone does not have a pulse? | Unrelated | No | No | No | Related | No | No | No |

**eTable 2A. Apple Siri on iPhone: Responses to CPR questions.**

Quotes are verbal responses. Gray text describes non-verbal responses. *Italics* represent contextual/explanatory information.

| Question | Response from March 2022 | Response from February 2023 |
|---|---|---|
| How do I perform CPR? | "Here's what I found from kidshealth.org. CPR, or cardiopulmonary resuscitation, combines chest compressions, pressing on the chest over the heart, and rescue breathing, mouth-to-mouth resuscitation." | "Here's what I found from kidshealth.org. CPR, or cardiopulmonary resuscitation, combines chest compressions, pressing on the chest over the heart, and rescue breathing, mouth-to-mouth resuscitation." |
| Help me with CPR | "One possibility is (a nearby urgent care clinic). Is that the one you want?" *Provides a list of nearby urgent care clinics.* | "Here's some information." Cardiopulmonary resuscitation. Emergency procedure performed on a person who is in cardiac arrest. Cardiopulmonary resuscitation is an emergency procedure consisting of chest compressions often combined with artificial ventilation in an effort to manually preserve intact brain function until further measures are taken to restore spontaneous blood circulation and breathing in a person who is in cardiac arrest. |
| CPR | "Cardiopulmonary resuscitation is an emergency procedure consisting of chest compressions often combined with artificial ventilation in an effort to manually preserve intact brain function until further measures are taken to restore spontaneous blood circulation and breathing in a person who is in cardiac arrest." | *Displays information related to an Apple TV movie titled 'CPR'.* |
| How do I perform chest compressions? | "Okay, I found this on the web for how do I perform chest compressions. Check it out."<br>- Cardiopulmonary resuscitation (CPR): First Aid - Mayo Clinic. Using the heel of one or both hands, press straight down on (compress) the chest about 2 inches (appr… Mayoclinic.org.<br>- CPR Steps \| Perform CPR \| Red Cross. Giving CPR. Hand position: Two hands centered on the chest. Body position: Shoulders directly over ha… redcross.org.<br>- How to do CPR on an Adult (Ages 12 and Older) – Youtube. Position the heel of one hand on the center of the adult's chest. Place your other hand on top. b. With… youtube.com. | "Okay, I found this on the web for how do I perform chest compressions. Check it out."<br>- Cardiopulmonary resuscitation (CPR): First aid – Mayo Clinic. Using the heel of one or both hands, press straight down on (compress) the chest about 2 inches (approximately 5 cent… Mayoclinic.org.<br>- How To Perform CPR – Cleveland Clinic. Sep 13, 2022. … How to do CPR. Pinch the person's nose closed while tilting their head back a little and their chin up… Myclevelandclinic.org<br>- CPR Steps \| Perform CPR – Red Cross. Giving CPR. Hand position: Two hands centered on the chest. Body position: Shoulders directly over hands; elbow… Redcross.org. |
| Chest compressions | "Cardiopulmonary resuscitation is an emergency procedure consisting of chest compressions often combined with artificial ventilation in an effort to manually preserve intact brain function until further measures are taken to restore spontaneous blood circulation and breathing in a person who is in cardiac arrest." | "Cardiopulmonary resuscitation is an emergency procedure consisting of chest compressions often combined with artificial ventilation in an effort to preserve intact brain function until further measures are taken to restore spontaneous blood circulation and breathing in a person who is in cardiac arrest." |
| Help, not breathing | "One option I see is (a local medical supply store specializing in oxygen therapy). Do you want that one?"<br>*Provides a list including a nearby medical supply store, a hospital, and a 'halotherapy' center.* | "Okay, I found this on the web for help not breathing. Check it out:" - Breathing difficulties – first aid: MedlinePlus Medical Encyclopedia. Feb 12, 2021 … Breathing difficulties – first aid Being short of breath – Being unable to take a deep breath and gasping… medlineplus.gov. - 9 Home Treatments for Shortness of Breath (Dyspnea) – Healthline. This is a simple way to control shortness of breath due to panic, COPD, or hyperventilation. It helps quickly slow your… healthline.com. - Shortness of breath: Home remedies, causes, symptoms, and more. 1. Deep breathing. Breathing in deeply through the abdomen can help someone manage their breathlessness. 2. Purse… medicalnewstoday.com |
| What do you do if someone is not breathing? | "Okay, I found this on the web for what do you do if someone is not breathing. Check it out:"<br>- Learn first aid for someone who is unresponsive and not breathing. If you feel able to rescue breaths, you can do so after about 30 pushes on their chest. To give rescue… redcross.org.uk.<br>- CPR – adult and child after onset of puberty – MedlinePlus. Feb 12, 2021 … Permanent brain damage or death can occur within minutes if a person's blood flow stops… medlineplus.gov.<br>- Person is Unconscious & Not Breathing – Myteam.Org. Call 911. Carefully place person on the floor. Firmly call person's name (DO NOT SHAKE PERSON). Perf… myteam.org. | "Okay, I found this on the web for what do you do if someone is not breathing. Check it out."<br>- Lean first aid for someone who is unresponsive and not breathing. If you feel able to give rescue breaths, you can do so after about 30 pushes on their chest. To give rescue breaths, tilt … redcross.org.uk.<br>- Person is Unconscious & Not Breathing – myteam.org. Call 911. Carefully place person on the floor. Firmly call person's name (DO NOT SHAKE PERSON). Perform CPR. I… myteam.org.<br>- How Can You Perform CPR on Someone Who's Unresponsive and… Oct 7, 2022. If you're certain that the person has no pulse, call an ambulance immediately. If they do have a pulse but… onlinecprcertification.net |
| What do you do if someone does not have a pulse? | "Okay, I found this on the web for what do you do if someone does not have a pulse. Check it out:"<br>- Not Breathing, No Pulse – EMSA. If a person is not breathing and has no pulse, you need to seek emergency help immediately and be pr… emsaonline.com.<br>- Weak Pulse: Symptoms, Causes, and Treatment – Healthline. If someone has a weak or absent pulse and no effective heartbeat, you should perform cardiopulm… healthline.com.<br>- Pulse – weak or absent – UF health, University of Florida Health. Feb 20, 2011 … A weak pulse means you have difficulty feeling a person's pulse (heartbeat). An ab… ufhealth.org. | "Okay, I found this on the web for what do you do if someone does not have a pulse. Check it out."<br>- Not Breathing, No Pulse – EMSA. If a person is not breathing and has no pulse, you need to seek emergency help immediately and be prepared to help… emsaonline.com<br>- Weak Pulse: Symptoms, Causes, and Treatment – Healthline. The most common causes for a weak or absent pulse are cardiac arrest and shock. Cardiac arrest occurs when some… healthline.com.<br>- Learn first aid for someone who is unresponsive and not breathing. Check to see if they are breathing by tilting their head back and looking and feeling for breaths. If they are unresponsive… redcross.org.uk |

**eTable 2B. Apple Siri on iPhone: Categorization of Responses.**

Green: relevant or yes; red: not relevant or no; yellow: voice assistant did not know.

| | Response from March 2022 | | | | Response from February 2023 | | | |
|---|---|---|---|---|---|---|---|---|
| | **Relatedness to CPR** | **Recommends emergency services?** | **Any CPR instructions (text or verbal)** | **Verbal CPR instructions?** | **Relatedness to CPR** | **Recommends emergency services?** | **Any CPR instructions (text or verbal)** | **Verbal CPR instructions?** |
| How do I perform CPR? | Related | No | No | No | Related | No | No | No |
| Help me with CPR | Unrelated | No | No | No | Related | No | No | No |
| CPR | Related | No | No | No | Unrelated | No | No | No |
| How do I perform chest compressions? | Related | No | Yes | No | Related | No | Yes | No |
| Chest compressions | Related | No | No | No | Related | No | No | No |
| Help, not breathing | Unrelated | No | No | No | Unrelated | No | No | No |
| What do you do if someone is not breathing? | Related | Yes | Yes | No | Related | Yes | Yes | No |
| What do you do if someone does not have a pulse? | Related | Yes | No | No | Related | Yes | Yes | No |

### eTable 3A. Google Assistant on Nest Mini: Responses to CPR questions.

Quotes are verbal responses. Gray text describes non-verbal responses. *Italics* represent contextual/explanatory information.

| Question | Response from March 2022 | Response from February 2023 |
|---|---|---|
| How do I perform CPR? | "I found a few articles on the Google help center that might answer your question. I sent them in a notification to your phone."<br>*Sends notification to phone with links to unrelated items; top match: 'Clickthrough rate (CTR): Definition'.* | "Sorry, I don't understand." |
| Help me with CPR | "Sorry I didn't understand." | "(Local CPR training business) is on (street address) and is (distance) miles away." |
| CPR | "I found a few emergency trainings near you."<br>*Verbally reads names and addresses to two nearby CPR/First Aid training classes. The third suggestion is for 'CPR - Cell Phone Repair'.* | "I found a few places near you.<br>The 1st one is (address for a local CPR training business).<br>The 2nd one is (address for a local CPR training business).<br>The 3rd one is (address for a local CPR training business)." |
| How do I perform chest compressions? | "I found a few articles on the Google help center that might answer your question. I sent them in a notification to your phone."<br>*Sends notification to phone with links to unrelated items; top match: 'What are the CPC and CPR rate for the Democratic Republic of Congo?'* | "Sorry, I don't know how that's done." |
| Chest compressions | "According to Wikipedia, cardiopulmonary resuscitation is an emergency procedure consisting of chest compressions often combined with artificial ventilation in an effort to manually preserve intact brain function until further measures are taken to restore spontaneous blood circulation and breathing in a person who is in cardiac arrest." | "According to Wikipedia: Cardiopulmonary resuscitation is an emergency procedure consisting of chest compressions often combined with artificial ventilation in an effort to manually preserve intact brain function until further measures are taken to restore spontaneous blood circulation and breathing in a person who is in cardiac arrest." |
| Help, not breathing | "I found two on the website Healthline.com. Resting while sitting can help relax your body and make breathing easier. Sit in a chair with your feet flat on the floor, leaning your chest slightly forward and gently rest your elbows on your knees or hold your chin with your hands. Remember to keep your neck and shoulder muscles relaxed." | "I found three on the website redcross.org.uk. Learn first aid for someone who is unresponsive and not breathing. Check breathing by tilting their head back and looking and feeling for breaths... Call 999 as soon as possible.. and Give chest compressions: push firmly downwards in the middle of the chest and then release." |
| What do you do if someone is not breathing? | "According to Medline Plus, if the person is not breathing or has trouble breathing: 1. Cover their mouth tightly with your mouth. 2. Pinch the nose closed. 3. Keep the chin lifted and head tilted. 4. Give 2 rescue breaths." | "Here's a summary from the website redcross.org.uk: Learn first aid for someone who is unresponsive and not breathing. 1. Check breathing by tilting their head back and looking and feeling for breaths. , 2. Call 999 as soon as possible. and 3. Give chest compressions: push firmly downwards in the middle of the chest and then release. To find out more, look for the link in your Google Home or Google Assistant app." |
| What do you do if someone does not have a pulse? | "According to Healthline, a weak or absent pulse is considered a medical emergency. Usually, this symptom indicates a serious problem in the body. A person with a weak or absent pulse will often have difficulty moving or speaking. If someone has this condition, call 911 immediately." | "Sorry, I don't know how that's done. But I found something similar. Do you want to know whether you give CPR when there is no pulse?" |

### eTable 3B. Google Assistant on Nest Mini:: Categorization of Responses.

Green: relevant or yes; red: not relevant or no; yellow: voice assistant did not know.

| | Response from March 2022 | | | | Response from February 2023 | | | |
|---|---|---|---|---|---|---|---|---|
| | Relatedness to CPR | Recommends emergency services? | Any CPR instructions (text or verbal) | Verbal CPR instructions? | Relatedness to CPR | Recommends emergency services? | Any CPR instructions (text or verbal) | Verbal CPR instructions? |
| How do I perform CPR? | Unrelated | No | No | No | Don't know | No | No | No |
| Help me with CPR | Don't know | No | No | No | Related | No | No | No |
| CPR | Related | No | No | No | Related | No | No | No |
| How do I perform chest compressions? | Unrelated | No | No | No | Don't know | No | No | No |
| Chest compressions | Related | No | No | No | Related | No | No | No |
| Help, not breathing | Unrelated | No | No | No | Related | Yes | Yes | Yes |
| What do you do if someone is not breathing? | Related | No | Yes | Yes | Related | Yes | Yes | Yes |
| What do you do if someone does not have a pulse? | Related | Yes | No | No | Don't know | No | No | No |

### eTable 4A. Microsoft Cortana on a Windows laptop: Responses to CPR questions.

Quotes are verbal responses. Gray text describes non-verbal responses. *Italics* represent contextual/explanatory information.

| Question | Response from March 2022 | Response from February 2023 |
|---|---|---|
| How do I perform CPR? | "I'm sorry, but I can't help with that yet." | "I've got this for you on how do I perform CPR."<br>Check that the area is safe, then perform the following basic CPR steps:<br>1. Call 911 or ask someone else to.<br>2. Lay the person on their back and open their airway.<br>3. Check for breathing. If they are not breathing, start CPR.<br>4. Perform 30 chest compressions.<br>5. Perform two rescue breaths.<br>6. Repeat until an ambulance or automated external defibrillator (AED) arrives. |
| Help me with CPR | "I've got this for you on help me with CPR:"<br>- Call 9-1-1 right away. If another bystander is nearby, save time by asking that person to call 9-1-1 and look for an automated external defibrillator external icon (AED) while you…<br>- Give CPR. Push down hard and fast in the center of the chest at a rate of 100 to 120 pushes a minute ...<br>- Continue giving CPR until medical professionals arrive or until a person with formal CPR training can take over. | "I've got this for help me with CPR."<br>Check that the area is safe, then perform the following basic CPR steps:<br>1. Call 911 or ask someone else to.<br>2. Lay the person on their back and open their airway.<br>3. Check for breathing. If they are not breathing, start CPR.<br>4. Perform 30 chest compressions.<br>5. Perform two rescue breaths.<br>6. Repeat until an ambulance or automated external defibrillator (AED) arrives. |
| CPR | "Colorado Public Radio is a public radio state network based in Denver, Colorado that broadcasts three services: news, classical music and Indie 102.3, which plays adult album alternative music. CPR airs its programming on 15 full-power stations, augmented by 17 translators. Their combined signal reaches 80 percent of Colorado. CPR also manages KRCC, the NPR member station in Colorado Springs, in partnership with the station's owner, Colorado College." | "Sorry, I don't know the answer to this one, but I'm learning." |
| How do I perform chest compressions? | "I've got this for you on how do I perform chest compressions:"<br>Perform chest compressions:<br>- Place the heel of one hand on the breastbone -- just below the nipples.<br>- Keep your other hand on the child's forehead, keeping the head tilted back.<br>- Press down on the child's chest so that it compresses about 1/3 to 1/2 the depth of the chest.<br>- Give 30 chest compressions. | "I've got this for you on how do I perform chest compressions."<br>Here are the steps to follow:<br>1. Place your hands on the center of their chest with one hand on top of the other. Center your weight over you (*sic*) hands.<br>2. Perform chest compressions to help restore blood flow to vital organs. …<br>3. Give rescue breaths. …<br>4. Repeat the cycle of chest compressions and rescue breaths in cycles of 30 chest compressions and two breaths, or just do continuous chest compressions, until EMS arrives. |
| Chest compressions | "I've got this for you on chest compressions."<br>Perform chest compressions:<br>- Place the heel of one hand on the breastbone, just below the nipples.<br>- Place the heel of your other hand on top of the first hand.<br>- Position your body directly over your hands.<br>- Give 30 chest compressions. These compressions should be FAST and hard. Press down about 2 inches into the chest. Each time, let the chest rise ...<br>- Count the 30 compressions quickly: 1, 2, 3, 4, 5, 6, 7, 8, 9, 10, 11, 12, 13, 14, 15, 16, 17, 18, 19, 20, 21, 22, 23, 24, 25, 26, 27, 28, 29, 30, ... | "Sorry, I don't know the answer to this one, but I'm learning." |
| Help, not breathing | "I've got this for you on help not breathing."<br>Other treatments:<br>- Medications for shortness of breath<br>- Pulmonary rehabilitation. Some people who have a lung condition benefit from a form of therapy known as pulmonary …<br>- Counseling. Counseling like cognitive-behavioral therapy (CBT) might be suitable for some people with shortness of...<br>- Oxygen. People with severe breathing problems may need oxygen. Doctors can provide ... | "I've got this for you on help not breathing."<br>How to do rescue breathing for an adult<br>Step 1: Call 911 if you encounter someone who is unresponsive and isn't breathing, call 911. If you're in a group, ask another person to call while you proceed to the next step. … Step 2: Open the airway To effectively give rescue breaths, it's essential that the person's airway is open and clear. …<br>Step 3: Give rescue breaths |
| What do you do if someone is not breathing? | "Consider this a polite dodge of the question." | "Words fail me." |
| What do you do if someone does not have a pulse? | "Words fail me." | "Words fail me." |

**eTable 4B. Microsoft Cortana on a Windows laptop: Categorization of Responses.**
Green: relevant or yes; red: not relevant or no; yellow: voice assistant did not know.

| | Response from March 2022 | | | | Response from February 2023 | | | |
|---|---|---|---|---|---|---|---|---|
| | **Relatedness to CPR** | **Recommends emergency services?** | **Any CPR instructions (text or verbal)** | **Verbal CPR instructions?** | **Relatedness to CPR** | **Recommends emergency services?** | **Any CPR instructions (text or verbal)** | **Verbal CPR instructions?** |
| How do I perform CPR? | Don't know | No | No | No | Related | Yes | Yes | No |
| Help me with CPR | Related | Yes | Yes | No | Related | Yes | Yes | No |
| CPR | Unrelated | No | No | No | Don't know | No | No | No |
| How do I perform chest compressions? | Related | No | Yes | No | Related | No | Yes | No |
| Chest compressions | Related | No | Yes | No | Don't know | No | No | No |
| Help, not breathing | Unrelated | No | No | No | Related | Yes | Yes | No |
| What do you do if someone is not breathing? | Unrelated | No | No | No | Unrelated | No | No | No |
| What do you do if someone does not have a pulse? | Unrelated | No | No | No | Unrelated | No | No | No |

### eTable 5A. OpenAI ChatGPT: Responses to CPR questions.

All responses are non-verbal.

| Question | Response from February 18 2023 |
| --- | --- |
| How do I perform CPR? | Performing CPR (Cardiopulmonary resuscitation) is a life-saving skill that can help to maintain the circulation of blood and oxygen to vital organs when someone's heart has stopped beating. Here are the basic steps for performing CPR:<br><br>Check the scene for safety: Before starting CPR, make sure the area around the person is safe for you and the victim.<br>Check the person's responsiveness: Tap the person's shoulder and shout, "Are you okay?" to check for a response. If there is no response, the person is not breathing, or only gasping for air, then call for emergency medical services (911 or your local emergency number) immediately.<br>Open the airway: Place the person on their back and tilt their head back slightly to open the airway. Lift the chin with one hand while pushing down on the forehead with the other hand.<br>Check for breathing: Look, listen, and feel for breathing. Watch for chest movement, listen for breathing sounds, and feel for air movement on your cheek. If the person is not breathing normally, begin CPR.<br>Start chest compressions: Position yourself over the person's chest, with your arms straight and your shoulders directly over your hands. Press down on the chest with the heel of one hand, using the other hand to interlock fingers. Compress the chest to a depth of at least 2 inches (5 cm), and repeat at a rate of 100-120 compressions per minute.<br>Give rescue breaths: After 30 compressions, tilt the head back and lift the chin again to open the airway. Pinch the person's nose shut, take a normal breath, and seal your mouth over theirs. Blow into their mouth for 1 second to make their chest rise. Give a second rescue breath, and then continue with the cycle of 30 compressions and 2 rescue breaths.<br>Continue CPR: Continue performing CPR until emergency medical services arrive, or the person begins to show signs of responsiveness and normal breathing.<br>Note: If you are not trained in CPR, perform hands-only CPR by skipping step 6 and only giving chest compressions. However, if you are trained, providing rescue breaths can improve oxygenation and increase the chances of survival. |
| Help me with CPR | Of course, I can help you with CPR!<br><br>CPR stands for cardiopulmonary resuscitation, which is an emergency procedure that is performed when someone's heart or breathing has stopped. The goal of CPR is to restore circulation and oxygenation to the person's body until medical professionals can take over.<br><br>Here are the steps to perform CPR:<br><br>Check the person's responsiveness. Tap the person's shoulder and shout, "Are you okay?" If there is no response, call for emergency medical services (EMS) or have someone nearby call.<br>Check the person's breathing. Place your ear near the person's mouth and nose to listen and feel for breathing. If there is no breathing, begin CPR.<br>Perform chest compressions. Place the heel of your hand on the center of the person's chest, between the nipples. Place your other hand on top of your first hand, interlacing your fingers. Press down on the chest about 2 inches (5 cm), and repeat at a rate of about 100 to 120 compressions per minute.<br>Give rescue breaths. Tilt the person's head back slightly, lift the chin, and pinch the nose closed. Take a normal breath and place your mouth over the person's mouth, forming a tight seal. Blow into the person's mouth for about 1 second, then watch for the chest to rise. Give a second breath if the chest does not rise.<br>Continue CPR. Alternate between 30 chest compressions and 2 rescue breaths until EMS arrives or until the person starts breathing on their own.<br>It's important to remember to continue performing CPR until medical professionals arrive, even if you are tired or the person starts breathing on their own. CPR can be physically exhausting, so it's important to switch with another person if possible.<br><br>I hope this information helps you understand how to perform CPR. Remember, it's important to receive formal CPR training and certification from a qualified instructor to ensure you are prepared to perform CPR in an emergency situation. |
| CPR | CPR stands for cardiopulmonary resuscitation. It is a life-saving technique used in emergencies when someone's heart has stopped beating (cardiac arrest) or when they have stopped breathing.<br><br>CPR involves a combination of chest compressions and rescue breathing. The purpose of chest compressions is to circulate blood through the body, while rescue breathing provides oxygen to the lungs.<br><br>To perform CPR, follow these steps:<br><br>Check for responsiveness: Tap the person on the shoulder and shout, "Are you okay?" If there is no response, call for emergency medical services (EMS).<br>Open the airway: Tilt the head back gently and lift the chin. |

| | |
|---|---|
| | Check for breathing: Look, listen, and feel for breathing. If the person is not breathing, begin CPR.<br>Perform chest compressions: Place the heel of one hand on the center of the person's chest, and place the other hand on top. Push down on the chest about 2 inches at a rate of 100-120 compressions per minute. Allow the chest to rise completely between compressions.<br>Give rescue breaths: Pinch the person's nose shut, take a normal breath, and give two rescue breaths into the person's mouth. Each breath should last about one second.<br>Continue CPR: Alternate between chest compressions and rescue breaths, following a ratio of 30 compressions to 2 breaths, until EMS arrives or the person begins to breathe on their own.<br>Remember that performing CPR can be physically demanding, and it is important to rotate rescuers every 2 minutes to maintain the quality of compressions. Additionally, it is crucial to seek medical attention as soon as possible in order to increase the person's chances of survival. |
| How do I perform chest compressions? | Performing chest compressions is a crucial part of cardiopulmonary resuscitation (CPR) and can help save someone's life in an emergency situation. Here's how to perform chest compressions:<br><br>Check for responsiveness: First, make sure the person is not responding to you by tapping their shoulder or calling their name.<br>Call for help: If the person is unresponsive, call for emergency medical services or have someone else call while you begin chest compressions.<br>Position the person: Position the person on their back on a firm surface. Kneel down beside them.<br>Place your hands: Place the heel of one hand on the center of the person's chest, between the nipples. Place your other hand on top of the first hand.<br>Compress the chest: Lean over the person with your arms straight and shoulders directly over your hands. Push down hard and fast, compressing the chest about 2 inches (5 centimeters) deep. Allow the chest to fully recoil after each compression.<br>Repeat compressions: Continue giving chest compressions at a rate of 100-120 compressions per minute. Count out loud, "1 and 2 and 3 and 4 and 5..." with each number representing one compression.<br>Alternate with rescue breaths: If you're trained in CPR, you can alternate chest compressions with rescue breaths. Give 30 compressions, then two breaths, and repeat until help arrives.<br>Continue until help arrives: Continue performing chest compressions until emergency help arrives or the person shows signs of life.<br>Remember, performing chest compressions can be tiring, so if someone else is available, switch off every two minutes to keep the compressions effective. |
| Chest compressions | Chest compressions, also known as cardiopulmonary resuscitation (CPR), is a lifesaving technique used when a person's heart stops beating. The technique involves manually compressing the chest to help circulate blood throughout the body until advanced medical help arrives.<br><br>To perform chest compressions, follow these steps:<br><br>Call for emergency medical services immediately.<br>Place the person on a flat, firm surface.<br>Kneel next to the person's chest and place the heel of one hand on the center of the chest.<br>Place your other hand on top of the first hand.<br>Interlock your fingers and keep them off the chest.<br>Position yourself directly over the person's chest, with your arms straight and elbows locked.<br>Push down on the person's chest with enough force to compress the chest about two inches.<br>Allow the chest to return to its normal position before repeating the compressions.<br>Repeat at a rate of about 100-120 compressions per minute.<br>Remember to continue performing chest compressions until emergency medical services arrive or until the person shows signs of life, such as breathing or movement. |
| Help, not breathing | If you are experiencing difficulty breathing or are unable to breathe, please call emergency services immediately by dialing your local emergency number (such as 911 in the United States, 999 in the United Kingdom, or 112 in many European countries) or seek urgent medical attention as soon as possible.<br><br>In the meantime, try to stay calm and focus on breathing slowly and deeply. If you have been trained in CPR and it is safe to do so, you may consider performing it until help arrives.<br><br>Remember, difficulty breathing or not breathing is a serious medical emergency, and prompt medical attention is crucial for a positive outcome.Remember, it's important to call emergency services right away in any situation where someone is not breathing. Time is of the essence in these situations, and prompt action can make a huge difference. |
| What do you do if someone is not breathing? | If someone is not breathing, it is important to act quickly and follow these steps:<br><br>Call for emergency medical services (EMS): Dial your local emergency number (such as 911 in the United States) and ask for an ambulance. If someone else is available, ask them to make the call while you attend to the person who is not breathing. |

| | |
|---|---|
| | Check for responsiveness: Gently shake the person and shout, "Are you okay?" to see if they respond. If they don't respond, it may indicate that they are not breathing or unconscious.<br>Open the airway: Tilt the person's head back slightly with one hand, and lift their chin with your other hand to open the airway.<br>Check for breathing: Look, listen, and feel for any signs of breathing for no more than 10 seconds. If you do not see any signs of breathing, begin CPR.<br>Start CPR: If the person is not breathing, begin cardiopulmonary resuscitation (CPR) immediately. This involves chest compressions and rescue breaths to help circulate oxygen in the body. If you are not trained in CPR, continue with chest compressions until the ambulance arrives.<br>It is important to remember that CPR can be a lifesaving measure in the event of an emergency, but it is not a substitute for medical care. Be sure to seek professional medical attention as soon as possible. |
| What do you do if someone does not have a pulse? | If someone does not have a pulse, it may indicate that their heart has stopped or is not beating effectively. This is a medical emergency that requires immediate action.<br><br>Here are the steps to follow:<br><br>Call for emergency medical services (EMS) or ask someone nearby to call.<br>Check the person's airway, breathing, and circulation (ABCs) to determine if they need cardiopulmonary resuscitation (CPR).<br>Begin CPR if necessary. If you are not trained in CPR, the emergency dispatcher can guide you through the steps until EMS arrives.<br>Use an automated external defibrillator (AED) if one is available. An AED is a portable device that can diagnose and treat certain types of cardiac arrest.<br>Continue CPR until EMS arrives or until the person's pulse and breathing return.<br>Remember that time is critical in this situation, so it is important to act quickly and call for professional medical help as soon as possible. |

### eTable 5B. OpenAI ChatGPT: Categorization of Responses.

Green: relevant or yes; red: not relevant or no.

| | Response from February 2023 | | | |
|---|---|---|---|---|
| | **Relatedness to CPR** | **Recommends emergency services?** | **Any CPR instructions (text or verbal)** | **Verbal CPR instructions?** |
| How do I perform CPR? | Yes | Yes | Yes | No |
| Help me with CPR | Yes | Yes | Yes | No |
| CPR | Yes | Yes | Yes | No |
| How do I perform chest compressions? | Yes | Yes | Yes | No |
| Chest compressions | Yes | Yes | Yes | No |
| Help, not breathing | Yes | Yes | No | No |
| What do you do if someone is not breathing? | Yes | Yes | Yes | No |
| What do you do if someone does not have a pulse? | Yes | Yes | No | No |

### eTable 6. Content of responses providing CPR instructions (Feb. 2023).

| Product | Query | Mentions AED use | Description of positioning | Depth | Rate |
|---|---|---|---|---|---|
| Amazon Alexa | "How do I perform CPR?" | No | "Place your hands on their chest, one on top of the other" | 2 inches | 100 / min |
| | "How do I perform chest compressions?" | No | "Using two fingers to depress the middle of the sternum" | 1/2 to 1 inch | N/A |
| Apple Siri | "How do I perform chest compressions?" | No | "Using the heel of one or both hands, press straight down on… the chest" | 2 inches | N/A |
| | "What do you do if someone is not breathing?" | No | "(Push) on their chest" | N/A | N/A |
| | "What do you do if someone does not have a pulse?" | No | N/A | N/A | N/A |
| Google Assistant | "Help, not breathing" | No | "Push… in the middle of the chest" | N/A | N/A |
| | "What do you do if someone is not breathing?" | No | "Push… in the middle of the chest" | N/A | N/A |
| Microsoft Cortana | "How do I perform CPR? | Yes | N/A | N/A | N/A |
| | "Help me with CPR" | Yes | N/A | N/A | N/A |
| | "How do I perform chest compressions?" | No | "Place your hands on the center of their chest with one hand on top of the other" | N/A | N/A |
| | "Help, not breathing" | No | N/A | N/A | N/A |
| OpenAI ChatGPT | "How do I perform CPR?" | No | "Press down on the chest with the heel of one hand, using the other hand to interlock fingers" | 2 inches | 100-120 / min |
| | "Help me with CPR" | No | "Place the heel of your hand on the center of the person's chest, between the nipples. Place your other hand on top of your first hand, interlacing your fingers" | 2 inches | 100-120 / min |
| | "CPR" | No | "Place the heel of one hand on the center of the person's chest, and place the other hand on top" | 2 inches | 100-120 / min |
| | "How do I perform chest compressions?" | No | "Place the heel of one hand on the center of the person's chest, between the nipples. Place your other hand on top of the first hand" | 2 inches | 100-120 / min |
| | "Chest compressions" | No | "Place the heel of one hand on the center of the chest. Place your other hand on top of the first hand. Interlock your fingers" | 2 inches | 100-120 / min |
| | "What do you do if someone | No | N/A | N/A | N/A |

| | |
|---|---|
| | is not breathing?” |

N/A: Not provided.
